# Integrated Plasma and Urinary Cell-free DNA Profiling Enables Noninvasive Molecular Detection from Ta to T4 Bladder Cancer

**DOI:** 10.64898/2026.07.13.26357430

**Authors:** Anja Lisa Riediger, Isabella Schindler, Martina Heller, Johannes Huber, Holger Sültmann, Magdalena Görtz

## Abstract

**Background and Objective:** Due to the heterogeneity of bladder cancer, minimally invasive molecular profiling may improve tumor characterization at the time of diagnosis. We evaluated whether integrated genomic and fragmentomic profiling of plasma and urinary circulating tumor DNA (ctDNA) detects BC-derived signals for diagnosis and disease stratification across all tumor stages.

**Methods:** In this real-world cohort, 202 plasma and urine samples were obtained from 33 patients with non-muscle-invasive BC (NMIBC), mostly Ta tumors, and 15 patients with muscle-invasive BC (MIBC), as well as from 58 cancer-free controls. Low-coverage whole-genome sequencing was performed to assess ctDNA fragmentation, chromosomal instability and copy number variations. Matched tumor tissue was analyzed to evaluate concordance between liquid biopsy and tissue-derived molecular alterations.

**Key Findings and Limitations:** Complementary genomic and fragmentomic profiling of cfDNA achieved detection rates of 75.8% in NMIBC patients and 91.7% in MIBC patients with paired plasma and urine. Distinct differences were observed between MIBC, NMIBC and cancer-free controls, consistent with increasing ctDNA signals during disease progression. Tumor tissue analysis confirmed BC-associated molecular alterations. Limitations include the single-center design and limited sample size.

**Conclusions and Clinical Implications:** Multimodal profiling of plasma and urinary cfDNA enabled the detection of tumor-derived molecular signals for all bladder cancer stages, including early-stage disease. By integrating genomic and fragmentomic features, this minimally invasive approach provides molecular tumor characterization at the time of diagnosis and may support future risk-adapted diagnostic, therapeutic and surveillance strategies.

**Advancing Practice:** *What does this study add?:* Our study demonstrates the potential of integrating multiple biomarkers from two liquid biopsy sources for minimally-invasive bladder cancer (BC) detection. This strategy enhances diagnostic sensitivity and addresses the limitations of single-analyte approaches, making it particularly relevant for future diagnostic and risk stratification in early stages, where circulating tumor-derived DNA levels are low. Multimodal liquid biopsy could serve as a valuable non-invasive method to guide clinical decisions, including additional diagnostics such as imaging and cystoscopy, as well as subsequent therapy management such as adjuvant therapy and follow-up.

*Patient Summary:* In this study, we explored a liquid biopsy approach using blood and urine to detect bladder cancer and assess its aggressiveness. We found that combining different genomic and fragmentomic markers improved the identification of tumor signals, even in early-stage cases. In the future, such approaches could help doctors to better assess bladder cancer risk and to personalize treatment decisions.

## INTRODUCTION

Bladder cancer (BC) is a biologically heterogeneous malignancy ranging from low-grade, non-muscle-invasive bladder cancer (NMIBC) to muscle-invasive and metastatic disease^1^. It is one of the most resource-intensive cancers due to its high recurrence rate and aggressiveness^2^. Current clinical risk stratification relies on histopathological parameters, such as tumor stage and grade, which only partially capture the underlying molecular biology of recurrence and progression^1^.

The limitations of current detection and surveillance strategies are particularly evident in NMIBC. Although cystoscopy is the cornerstone of diagnosis and follow-up, it is invasive and burdensome for patients and healthcare systems^3^. Urine cytology has high specificity for high-grade disease but limited sensitivity, particularly for low-grade tumors^4^. Consequently, patients with indolent, low-grade Ta tumors may undergo repeated procedures despite a low risk of progression. Meanwhile, patients with biologically aggressive disease may not be identified early enough for timely treatment intensification^1^. In muscle-invasive bladder cancer (MIBC), adjuvant treatment decisions after cystectomy remain challenging because conventional pathological features do not reliably identify all patients with occult micrometastatic spread^5^.

Liquid biopsy is emerging as a promising strategy to address these unmet needs. Circulating tumor DNA (ctDNA) is tumor-derived cell-free DNA that carries somatic alterations, such as copy-number alterations and fragmentomic features^6^. In BC, plasma ctDNA may reflect systemic tumor burden and occult metastatic potential, whereas urinary tumor DNA is particularly sensitive due to the direct contact between tumor tissue and urine^6,7^.

Whole-genome sequencing of urine cfDNA enabled the sensitive detection of minimal residual disease and predicted survival after radical cystectomy^8^. The phase 3 IMvigor011 trial demonstrated that ctDNA can identify patients at high risk of recurrence after cystectomy and that ctDNA-guided adjuvant immunotherapy resulted in significantly longer disease-free survival than placebo^9^. Recent data in high-grade pT1 NMIBC suggest that plasma and urinary ctDNA can identify patients with poor survival. However, using the GALEAS Bladder mutation test, in their cohort of 93 patients with high-grade pT1 NMIBC only ten (11%) were found to be ctDNA-positive^10^.

These studies suggest that tumor DNA in plasma and urine could enable more personalized detection, surveillance, treatment intensification, or de-escalation. However, most studies have focused on high-risk BC or post-cystectomy molecular residual disease. Few studies have profiled BC across the full clinical spectrum^11^, from Ta to metastatic disease, using paired plasma and urine while integrating matched tumor tissue. Therefore, we performed genomic and fragmentomic profiling of plasma and urinary ctDNA and assessed concordance between liquid biopsy and tissue-derived molecular alterations. We hypothesize that multimodal plasma and urinary ctDNA profiling can detect tumor-derived alterations and reflect tissue tumor biology for diagnosis and risk-adapted follow-up.

## METHODS

The Supplementary Material provides detailed descriptions of all method sections.

### Patient cohort and biological samples

48 BC patients (33 with NMIBC and 15 with MIBC) and 58 cancer-free controls were recruited at the Urology Clinic at Heidelberg University Hospital (September 2024 – March 2026). Plasma was available for 47 BC patients, urine for 44 patients and matched tumor tissue for 21 patients. Blood and urine samples were collected prior to transurethral resection or radical cystectomy. When available, matched tumor tissue from transurethral resection was analyzed. The control cohort included men undergoing cancer screening, unclear hematuria, or treatment for benign urological conditions. All individuals provided informed consent and the study was approved by the ethics committee of the Medical Faculty of Heidelberg University (S-404/2024 and S-130/2021).

### Liquid biopsy analyses

Plasma, urinary and tissue DNA were extracted and analyzed by low-coverage whole-genome sequencing (lcWGS) following previously established protocols^12^. One tissue sample was discarded because it had fewer than 20M reads. Copy-number profiling with tumor fraction (TFx) estimation, chromosomal instability analysis (CIA), and fragmentomic profiling were performed. CtDNA positivity was assessed based on six characteristic LBx features with predefined ctDNA detection thresholds: TFx and CIA score in both plasma and urine, plasma 10-bp oscillation score, and urinary 163–169 bp fragment proportion.

### Statistical analyses

Continuous variables are reported as median and interquartile range. Categorical variables are reported as n (%). Statistical analyses were conducted in R (v4.4.3)^13^, and visualizations were generated using ggplot2^14^. Group comparisons used Wilcoxon or Kruskal-Wallis tests, followed by Dunńs post hoc test, with Benjamini-Hochberg correction.

## RESULTS

### Study cohort

In this real-world study, 48 patients with Ta-T4 BC and 58 cancer-free controls were included (**Table 1**). 33 patients had NMIBC, mostly Ta disease, while 15 had MIBC. LcWGS analysis was performed on 47 plasma, 44 urine and 20 tumor tissue samples from BC patients. 43 patients were recruited before TURB and 5 patients before radical cystectomy. 9/43 patients had a prior BC diagnosis. 58 cancer-free controls (10 females and 48 males) were included, with a median age of 58.4 (51.1-62.9) years.

**Table 1.**
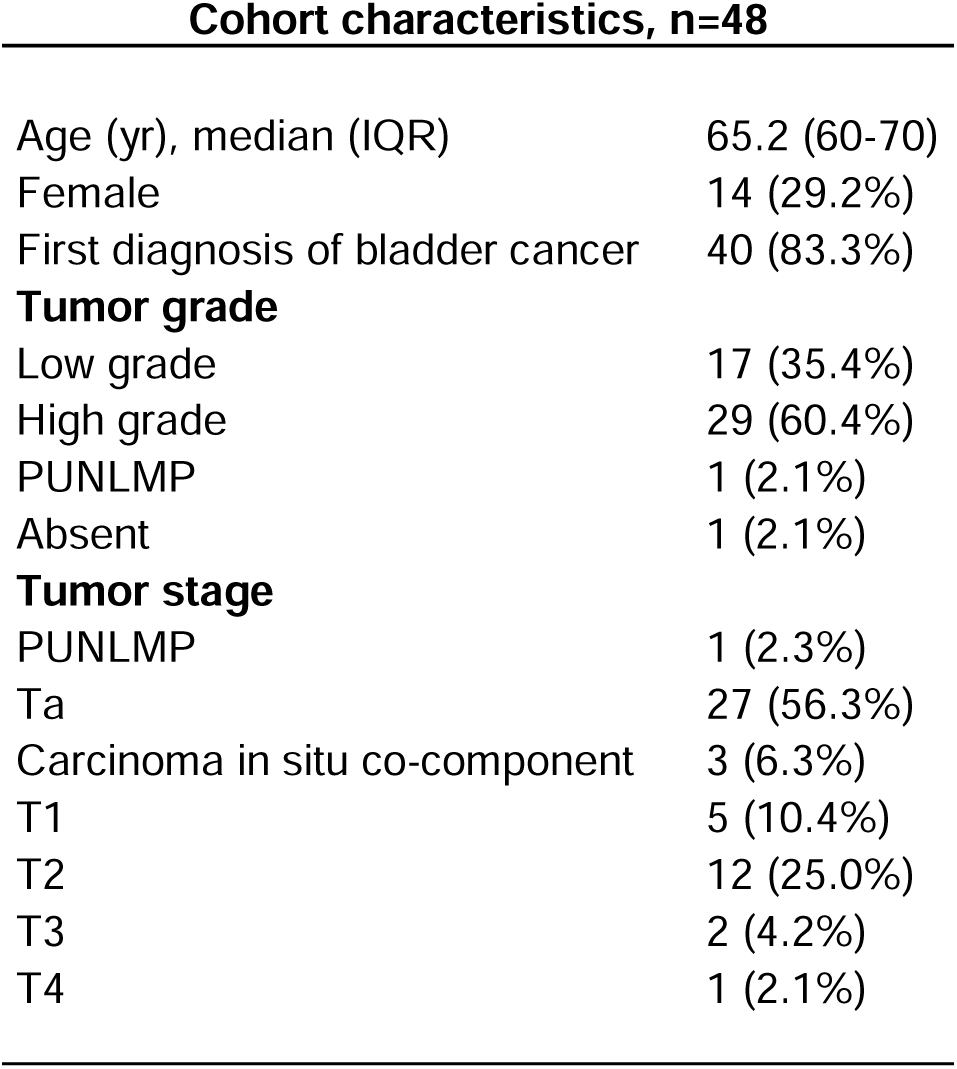
Clinical and Pathological Patient Characteristics of Bladder Cancer Cohort.

### Multimodal liquid biopsy detects tumor-derived signals across all bladder cancer stages

At least one positive LBx signal was detected in 75% (36/48) of BC patients using genomic and fragmentomic analyses across both biofluids. High detection rates were observed for all tumor stages. 75.8% of patients with NMIBC (25/33) and 73.3% of patients with MIBC (11/15) had at least one positive signal **(Fig. 1A**). As urine samples were missing for three MIBC patients, 91.7% (11/12) of MIBC patients with paired urine and plasma data were ctDNA-positive (*Supplementary Figure S1*). Thus, multimodal plasma and urinary ctDNA profiling identified tumor-derived molecular alterations in most BC patients at the time of diagnosis, including those with Ta BC. Single-parameter analyses yielded the highest ctDNA detection rates for genomic urinary cfDNA features, outperforming both genomic and fragmentomic plasma features **(Fig. 1A, B**). Plasma cfDNA features and urinary P163–169 showed comparable detection rates in NMIBC and MIBC, whereas urinary TFx and CIA scores achieved substantially higher detection rates in MIBC than in NMIBC (**Fig. 1B**). Combining plasma and urine analyses improved detection rates only slightly, reflecting the already high performance of urinary cfDNA markers. Integration of genomic and fragmentomic features across both biofluids further enhanced ctDNA detection, particularly in NMIBC (**Fig. 1A**). The burden of ctDNA-positive molecular features increased with tumor grade and tumor stage, with high-grade tumors exhibiting a higher number of ctDNA-positive features than low-grade tumors (**Fig. 1C** left) as well as advanced tumor stages exhibiting a higher number of ctDNA-positive features than low tumor stages (**Fig. 1C** right). Most patients with 0-1 ctDNA-positive results out of six analyzed features had low-grade BC, whereas all tumors with 4-6 positive features were high-grade BC. A similar pattern was seen for ctDNA-positive results and tumor stage.

**Figure 1.**
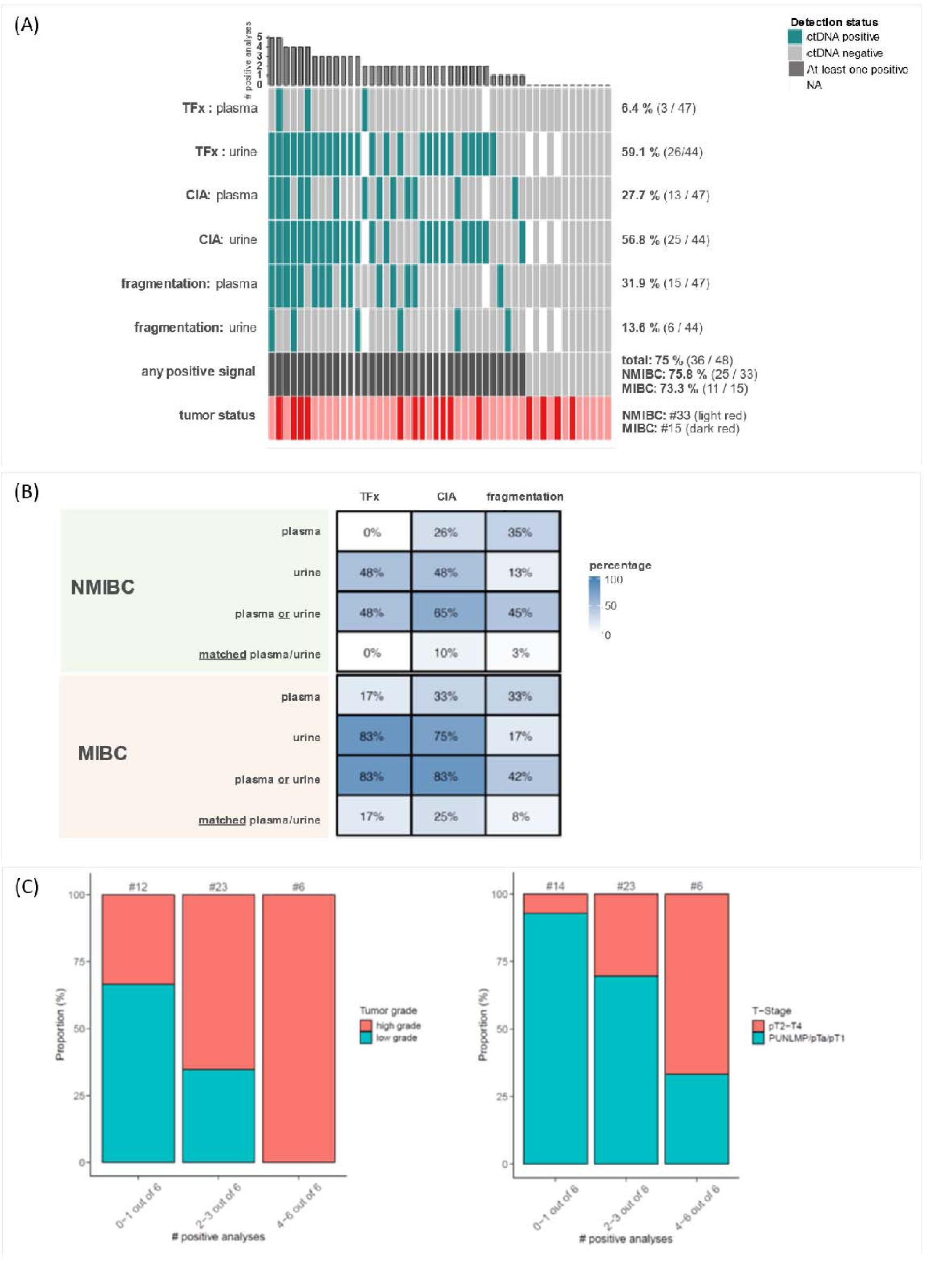
Multimodal ctDNA detection in plasma and urinary cfDNA from BC patients. **(A)** Oncoprint summarizing genomic (estimated TFx based on CNVs and CIA score) and fragmentomic (plasma 10-bp oscillation score, urinary P163–169 bp) analyses of plasma and urinary cfDNA from all 48 BC patients. Colored tiles indicate ctDNA-positive results in the respective analyses. White tiles represent unavailable LBx samples. The bar plot above the oncoprint shows the number of positive analyses per patient. The second-to-last row represents the number of BC patients with at least one ctDNA-positive result. **(B)** CtDNA detection rates in matched plasma and urine samples from NMIBC (top, n = 31) and MIBC (bottom, n = 12) patients with complete data sets (n = 43). Detection rates (percentages, %) are shown for individual plasma and urine analyses, matched plasma and urine samples, and the complementary plasma-urine approach combining results from both biofluids. **(C)** Association between the number of ctDNA-positive analyses and tumor grade (left) as well as between the number of ctDNA-positive analyses and tumor stage (right). Patients with matched plasma-urine pairs (n = 43) were grouped according to the number of positive analyses (0-1, 2-3, or 4-6 out of six) and tumor grade vs. tumor stage. Total patient numbers are shown above each bar. Tumor grade was unavailable for two patients. Abbreviations: cfDNA, cell-free DNA; CIA, chromosomal instability analysis; CNV, copy number variation; ctDNA, circulating tumor DNA; LBx, liquid biopsy; MIBC, muscle-invasive bladder cancer; NMIBC, non-muscle-invasive bladder cancer; P163–169 bp, proportion of cfDNA fragments with lengths of 163–169 bp; TFx, tumor fraction.

### Genomic profiling of plasma and urinary cfDNA identified tumor-associated characteristics in BC patients

Genome-wide copy number variations (CNVs) were profiled in plasma and urinary cfDNA via lcWGS, estimating TFx. CNVs were predominantly identified in urine for both NMIBC and MIBC, with increasing genomic burden in MIBC. In urine, BC patients demonstrated a significantly higher median TFx compared to controls (p = 7.13 e-09, *Supplementary Figure S2*). Stratification by tumor stage also revealed significant differences between controls and both MIBC and NMIBC (adjusted p = 7.27 e-06 and 5.40 e-06, respectively; **Fig. 2A** left). Plasma TFx values differed not significantly between BC patients and controls (p = 0.268, **Fig. 2A** right, *Supplementary Figure S2*). 26 urine samples (10x MIBC, 16x NMIBC) harbored TFx values above the ctDNA detection threshold. CNVs were also detected in matched plasma samples from two BC patients (one patient example in **Fig. 2B**). One additional patient had a ctDNA-positive plasma sample but lacked an available urine sample. Matched tumor tissue analysis showed that LBx-derived copy-number signals reflect tissue tumor biology. CNVs were identified in 18 out of 20 BC tissue samples from low to high extent. LBx samples with TFx >10% harbored recurrent genomic alterations, consistent with findings from the CNV analysis in BC tissue samples and known alterations in BC^15^. At the chromosome-arm level, recurrent deletions (1q, 2p, 3, 5p, 7p, 8q, 10p, 12, 17q, 19q, 20q, and 21q) and gains (2q, 4q, 5q, 6q, 8p, 9, 10q, 14q, 16q, 17p, and 22q) were detected in both tumor tissue and liquid biopsy samples, with frequencies of up to 50% (**Fig. 2C**). These findings support the biological validity of the LBx signals and suggest that they may serve as a noninvasive surrogate for tumor-derived genomic alterations in BC.The CIA score served as an additional genomic biomarker by focusing on regions with the greatest deviation from the copy-neutral state. BC samples had significantly higher median CIA scores than controls in both plasma (p = 1.35 e-09) and urine (p = 8.20 e-09). Stratification by tumor stage revealed significant differences between controls and both MIBC and NMIBC in plasma (adjusted p = 4.53 e-06 and 1.45 e-06, respectively; **Fig. 2D** left) and urine (adjusted p = 5.48 e-06 and 6.03 e-06, respectively; **Fig. 2D** right). 13 plasma (4x MIBC, 9x NMIBC) and 25 urine samples (7x MIBC, 18x NMIBC) exceeded the control-derived ctDNA detectability threshold.

**Figure 2:**
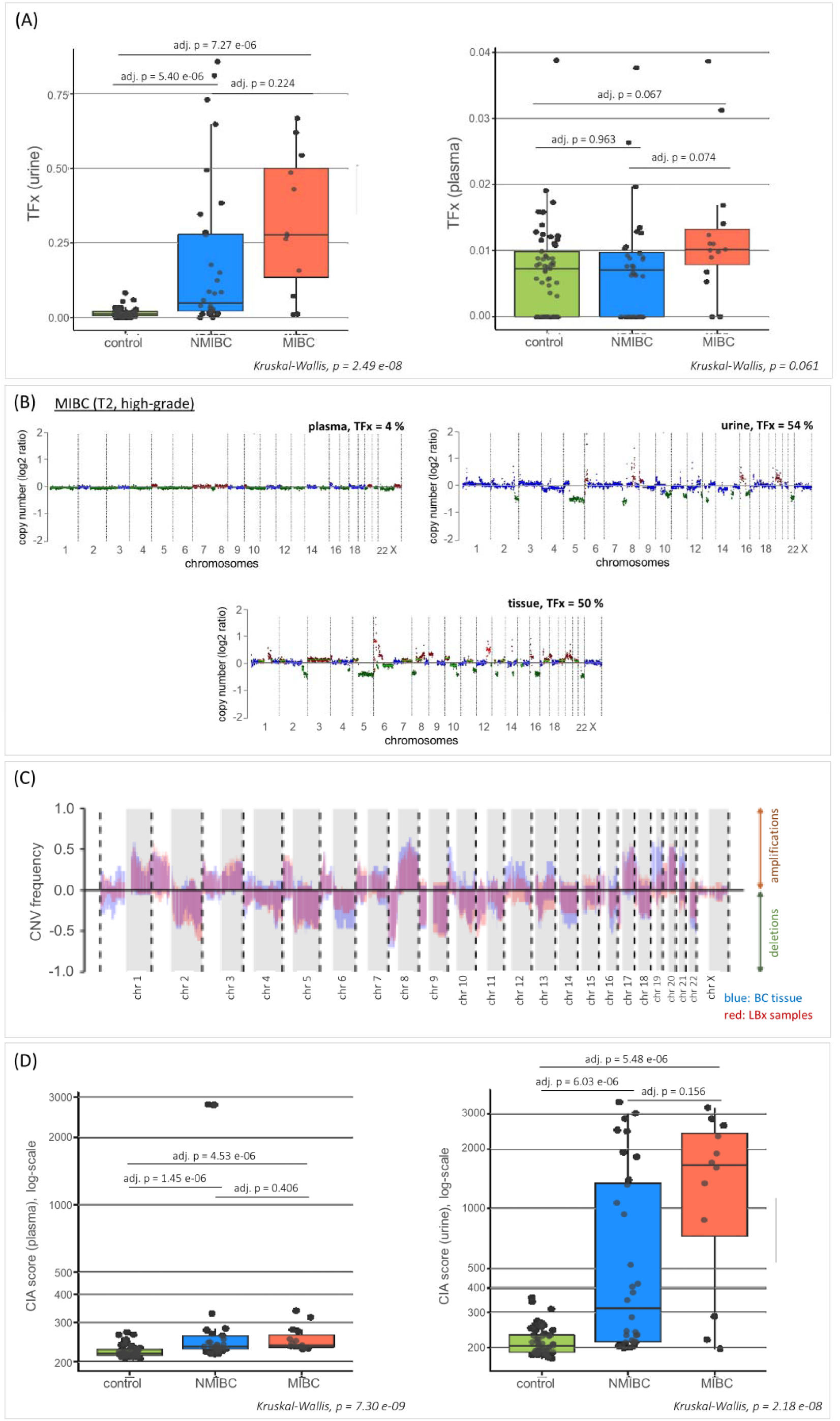
Genomic analyses in plasma and urinary cfDNA as well as correlation to BC tissue DNA. **(A)** Comparison of TFx values in plasma cfDNA (right) and urinary cfDNA (left) across cancer-free controls, NMIBC patients, and MIBC patients. **(B)** Complementary CNV profiles of plasma (left, top), urine (right, top) and BC tissue (bottom) samples from one MIBC patient. CNVs were detectable in all three sample types. **(C)** Summary of recurrent amplifications and deletions in BC LBx samples (red color) and tissue samples (blue color). Only BC LBx and tissue samples with detectable CNVs and an estimated TFx > 10% were considered (LBx: n = 21, tissue: n = 17). The y-axis indicates the frequency of a detected copy number state at the chromosomal coordinate specified on the xlJaxis across the samples. Areas shaded in gray represent the q-arm of the respective chromosome. **(D)** Comparison of CIA scores in plasma cfDNA (left) and urinary cfDNA (right) across cancer-free controls, NMIBC patients, and MIBC patients. **(A, D)** Box plot center lines indicate the median, and boxes illustrate the interquartile range with Tukey whiskers. Each dot represents one sample. Groups were compared using Kruskal-Wallis testing, followed by Dunn’s post hoc test. Abbreviations: chr, chromosome; cfDNA, cell-free DNA

### Plasma and urinary cfDNA fragmentation differed between BC patients and cancer-free controls

We analyzed global cfDNA fragmentation profiles and various fragmentation features (i.e., proportions and ratios of fragment length ranges, *Supplementary Figures S3+S4*) in BC patients and controls, to identify tumor-associated patterns and infer ctDNA presence. Plasma and urinary cfDNA fragmentation showed distinct characteristics and differed between BC patients and controls, as well as between NMIBC and MIBC. Plasma cfDNA fragmentation profiles exhibited a prominent 167 bp peak, representing nucleosome-associated DNA, with peak height increasing across controls, NMIBC and MIBC (**Fig. 3A**). Contrastingly, urinary cfDNA displayed broader fragment length distribution spanning 30-400 bp (**Fig. 3B**). Some samples contained an additional ∼167 bp peak, with increased extent in MIBC (**Fig. 3B**). Both biofluids exhibited prominent 10-bp oscillation patterns in fragments <150 bp, which were more pronounced and extended to longer fragments in urine (**Fig. 3A, B**).

**Figure 3:**
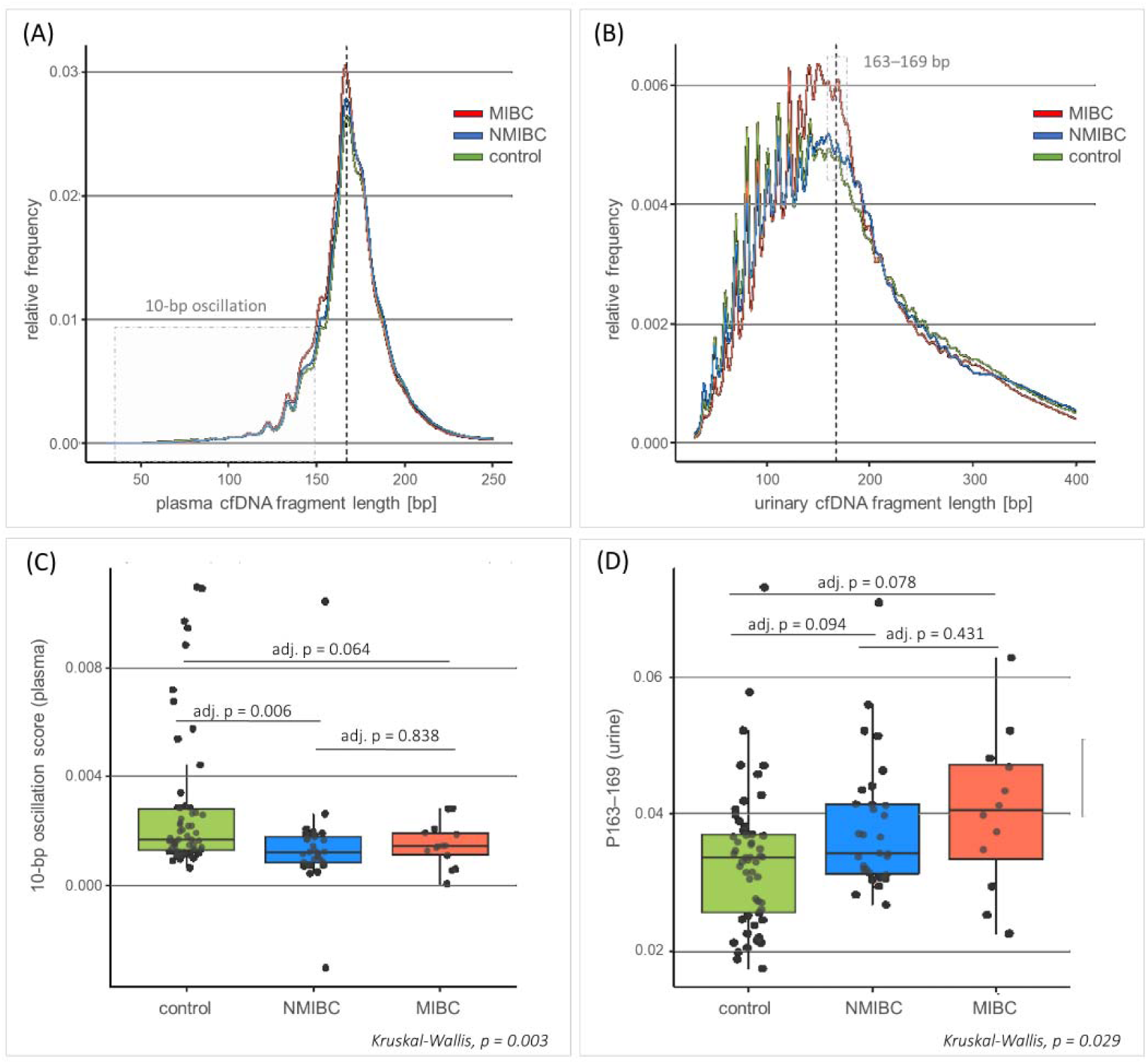
Fragmentation analysis of plasma and urinary cfDNA. **(A)** Median plasma cfDNA fragmentation profiles of cancer-free controls, NMIBC patients, and MIBC patients. The fragment length range of 30-150 bp is highlighted, illustrating the characteristic 10bp-oscillation pattern with local maxima and minima at 10-bp intervals. **(B)** Median urinary cfDNA fragmentation profiles of cancer-free controls, NMIBC patients, and MIBC patients. The fragment length range of 163-169 bp is highlighted. **(A+B)** Y-axis: Relative frequency of cfDNA fragments of a given length (bp) normalized to all fragments between 30 and 700 bp. The vertical gray dotted line indicates 167 bp. **(C)** Comparison of plasma 10-bp oscillation scores, calculated as the difference between the summed heights of local maxima and the summed depths of local minima, across cancer-free controls, NMIBC patients, and MIBC patients. **(D)** Comparison of urinary P163-169 bp across cancer-free controls, NMIBC patients, and MIBC patients. **(C+D)** Box plot center lines indicate the median, and boxes illustrate the interquartile range with Tukey whiskers. Each dot represents one sample. Groups were compared using Kruskal-Wallis testing, followed by Dunn’s post hoc test.

In plasma, BC patients harbored a significantly lower 10-bp oscillation score compared to controls (p = 3.93 e-05, adjusted p = 0.0003), although no consistent decline was observed across controls, NMIBC and MIBC patients (**Fig. 3C**). In urine, BC patients showed increased proportions of fragments with 163-169 bp length, especially for MIBC compared to controls (p = 0.026, adjusted p = 0.078), consistent with the additional peak at ∼167 bp (**Fig. 3D**). Using control-derived thresholds, analysis of these fragmentation features identified 15 BC patients (4x MIBC, 11 x NMIBC) with low plasma 10-bp oscillation scores and six BC patients (2x MIBC, 4x NMIBC) with high urinary P163-169 values, indicating ctDNA presence.

## DISCUSSION

In this real-world cohort, we conducted integrated genomic and fragmentomic profiling of plasma and urinary ctDNA at time of BC diagnosis spanning the full clinical spectrum from Ta NMIBC to metastatic MIBC. Our multimodal liquid biopsy approach detected tumor-derived molecular alterations in most patients, with complementary contributions from plasma and urine. Detection rates were high in NMIBC (75.8%), even in Ta tumors, and were particularly strong in MIBC when both biofluids were available, with 91.7% of patients being ctDNA-positive. Molecular signals detected in plasma and urinary ctDNA showed concordance with matched BC tissue. These results imply that a multi-biofluid, multi-feature liquid biopsy strategy can capture tumor-derived signals in both advanced disease and NMIBC, where tumor-derived cfDNA levels are low.

BC is among the most expensive malignancies to manage, with costs driven by late diagnosis, long-term surveillance, repeated procedures, radical surgery, and systemic therapies^2,16^. Noninvasive biomarkers may reduce costs by improving risk-adapted, biology-informed treatment strategies^4^. Recently, the International Bladder Cancer Group has emphasized deintensification strategies such as active surveillance, office fulguration, and chemoablation for carefully selected patients with recurrent low-grade Ta disease^17^. Our approach has several potential clinical applications, including diagnosis, surveillance intensity, and treatment selection^11^. For diagnosis, urine-based cfDNA profiling could complement cystoscopy and cytology, especially for patients with hematuria, high-risk exposures, or equivocal results. In surveillance, repeated urine-based molecular testing could support individualized follow-up strategies. The DaBlaCa-15 randomized controlled trial demonstrated that urinary biomarker-guided surveillance can reduce the need for cystoscopies without compromising short-term oncological safety, although the applied assay was not ctDNA-based^18^. For treatment strategies, plasma and urinary cfDNA profiling may help identifying high-risk patients who require intensified or adjuvant therapy strategies^5^.

In our study, urine detection was superior compared to plasma, which is biologically plausible due to the direct shedding of BC ctDNA into urine^6^. This is depicted by the higher CNV burden with predominantly detectable ctDNA in urine based on genomic analyses. Furthermore, an additional ∼167bp peak is visible in urinary cfDNA as sign of direct ctDNA shedding into urine, in contrast to glomerulo-filtrated cfDNA fragments which get further degraded during urinary tract passage^19^. However, plasma CIA and plasma fragmentation identified additional positive cases in our cohort. The high MIBC detection rate among patients with complete plasma-urine pairs aligns with the findings of Christensen et al., who detected tumor DNA in 89% of urine samples and 43% of plasma samples from MIBC patients, and showed that combined plasma and urine tumor DNA analysis has potential to predict treatment response and outcome^20^. In a previous study of high-grade pT1 NMIBC, ctDNA positivity was associated with worse overall and disease-specific survival^10^. A recent review of tumor DNA studies in BC concluded that plasma and urinary tumor DNA are promising tools for early detection, prognostic stratification, and therapeutic intensification^21^.

Our study has several strengths. First, the cohort reflects a real-world urological setting, includes both NMIBC and MIBC at the time of diagnosis, and evaluates ctDNA detection even in BC with Ta stage. Integrating genomic and fragmentomic features enabled broader detection than single features alone. Paired plasma and urine samples enabled direct comparison of biofluid-specific performance. Multimodal liquid biopsy features were derived from a single lcWGS assay, providing a broad, cost-efficient and scalable framework^22,23^ with potential for future integration into clinical diagnostics. Finally, matched tumor tissue samples supported the biological validation of liquid biopsy signals.

Several limitations need to be acknowledged. First, the sample size was limited. Second, this was an observational, single-center study and cannot establish clinical utility. Third, long-term recurrence data was unavailable. Fourth, urine samples were missing from three MIBC patients, which likely led to an underestimation of the MIBC detection rate. Fifth, tissue concordance may be affected by tumor heterogeneity and sampling sites.

Future studies should validate our findings in larger, prospective cohorts with standardized, paired plasma and urine collection. Serial sampling before and after TURB, intravesical therapy, radical cystectomy, and systemic therapy are required to determine whether molecular signal clearance or persistence have clinical value. Prospective trials are needed to evaluate whether multimodal, cfDNA-guided pathways can improve detection, reduce cystoscopies, and refine treatment strategies.

## Conclusion

Our study demonstrated that integrating genomic and fragmentomic profiling of plasma and urinary cfDNA improved diagnostic sensitivity across all BC tumor stages. Among NMIBC patients, 75.8% were ctDNA-positive, and among MIBC patients with paired plasma and urine data, 91.7% were ctDNA-positive. Urinary copy-number and chromosomal instability features were the strongest contributors, while plasma fragmentomics provided complementary information. These findings support multimodal plasma and urinary cfDNA profiling as a promising, noninvasive approach to detect tumor-derived molecular signals at the time of diagnosis. If validated prospectively, this approach could enable future risk-adapted diagnostic, treatment, and surveillance strategies.

**ABBREVIATIONS**
BC: Bladder cancer
cfDNA: Cell-free DNA
CIA: Chromosomal instability analysis
CNV: Copy number variations
ctDNA: Circulating tumor DNA
LBx: Liquid biopsy
lcWGS: Low-coverage whole genome sequencing
MIBC: Muscle invasive bladder cancer
NMIBC: Non-muscle invasive bladder cancer
P163-169 bp: Proportion of fragments with 163-169 bp length
TFx: Tumor fraction

## DECLARATIONS

### Ethical approval

The study was approved by the ethical committee of the University of Heidelberg (Approval No. S-404/2024 and S-130/2021) and was performed in accordance with the Declaration of Helsinki. Written informed consent was obtained from all participants prior to study inclusion.

### Contributorship

Conception and design: A.L.R., M.G., H.S.; Acquisition of data: A.L.R., I.S., M.H.; Analysis and interpretation of data: A.L.R., M.G., H.S.; Drafting of the manuscript: A.L.R., M.G., I.S.; Critical revision of the manuscript for important intellectual content: A.L.R., M.G., H.S., I.S.; Statistical analysis: A.L.R., M.G.; Obtaining funding: M.G.; Administrative, technical, or material support: M.G., H.S., I.S.; Supervision: M.G., H.S., J.H. All authors critically reviewed, edited, and added to the manuscript as well as approved the final version of the manuscript.

### Data availability statement

The data sets generated during and analyzed during the current study are available from the corresponding author on reasonable request.

### Declaration of generative AI and AI-assisted technologies in the writing process

During the preparation of this work the authors used ChatGPT-5 to search literature and to improve readability. After using this tool, the authors reviewed and edited the content as needed and take full responsibility for the content of the publication.

## Data Availability

All data produced in the present study are available upon reasonable request to the authors.

## Acknowledgements

The authors are grateful to Tommi Rantapero (Genevia Technologies) for substantial support with the bioinformatic analyses. In addition, we are grateful to the DKFZ NGS Core Facility for sequencing analyses and the DKFZ Omics IT and Data Management Core Facility for data management and processing. We sincerely thank the patients for their participation, as well as the medical staff at the Department of Urology at Heidelberg University Hospital for their support, especially Daniela Janscho.

## Take Home Message

Integrated genomic and fragmentomic profiling of plasma and urinary cell-free DNA improved tumor signal detection across all bladder cancer stages. Matched tissue analyses confirmed molecular alterations, supporting this minimally invasive approach for diagnosis, surveillance, and personalized treatment in bladder cancer.

## Supplementary Material

Integrated Plasma and Urinary Cell-free DNA Profiling Enables Noninvasive Molecular Detection from Ta to T4 Bladder Cancer Anja Lisa Riediger, Isabella Schindler, Martina Heller, Johannes Huber, Holger Sültmann, Magdalena Görtz

### Supplementary Methods

#### Patients

In this single-center study, patients at Heidelberg University Hospital were prospectively recruited between September 2024 and March 2026. 48 patients with histologically proven NMIBC or MIBC at diagnosis were retrospectively selected for this study. Inclusion criteria comprised availability of LBx samples and clinicopathologic data. LBx samples (blood and urine) were collected prior to TURB or radical cystectomy. Plasma and urinary cfDNA profiling were used to identify tumor-associated characteristics in patients with BC compared with cancer-free controls. 58 cancer-free patients represented the control cohort. Of these, plasma samples were available for 58 patients and urine samples for 53 patients. All individuals provided written informed consent, and the study was approved by the ethic committee of the Medical Faculty of Heidelberg University (S-404/2024 and S-130/2021).

#### Liquid biopsy analyses

Peripheral blood (2 x 9 ml) and urine samples (30-50 ml) were collected from participants. Plasma and urinary cfDNA isolation, library preparation and lcWGS were performed following previously established protocols^1^.

Sequencing reads were adapter trimmed, quality filtered and aligned to the human reference genome (hg19). LcWGS data were subsequently used for copy number profiling with tumor fraction (TFx) estimation, chromosomal instability analysis (CIA), and cfDNA fragmentation analyses. Genome-wide copy number analyses were performed using the ichorCNA algorithm^2^ with fluid type-matched panels of normals generated from control samples. CNV analyses were performed using the ichorCNA algorithm^2^ with default settings, except for the following parameter modifications: normal = “c(0.5, 0.6, 0.7, 0.8, 0.9, 0.95, 0.99, 0.995, 0.999, 1)”, maxCN = 3, estimateScPrevalence = FALSE. CIA scores were derived from normalized genome-wide copy number profiles by quantifying the most aberrant genomic bins (95^th^-99th percentile of absolute z-score values) relative to a copy-neutral state. Fragmentation analyses included genome-wide assessment of plasma and urinary cfDNA fragment-length distributions (30-700 bp), fragment-size interval proportions and ratios, as well as evaluation of the characteristic 10-bp oscillation pattern in short cfDNA fragments (30–150 bp), as previously described^1^.

Feature-specific ctDNA positivity thresholds were established based on the control cohort using percentile-based cutoffs (95th-percentile for plasma and urinary CIA scores, urinary TFx, and urinary P163-169 bp; 5th-percentile for plasma 10-bp oscillation score). As the 95th-percentile TFx value in plasma controls was below the ichorCNA limit of detection, a threshold of ≥0.03 was applied for plasma TFx in accordance with ichorCNA recommendations^2^. Tumor samples exceeding the predefined thresholds were classified as ctDNA-positive.

#### Bladder cancer tissue analyses

Histopathological assessment was conducted at the Institute of Pathology of Heidelberg University Hospital for all 48 patients. Fresh-frozen BC tissue samples were obtained from 21 patients (matching LBx samples were also analyzed) undergoing TURB at Heidelberg University Hospital. Extracted intraoperatively, the tissue samples (∼80 mg) were cryopreserved in liquid nitrogen and stored at −80°C until further processing. Genomic DNA was extracted from 21 fresh-frozen BC tissue samples. DNA isolation was performed with DNeasy Blood & Tissue Kit (Qiagen, Germantown, MD, USA), respectively, following the manufacturer’s guidelines. Sequencing libraries were prepared using the KAPA EvoPlus V2 Kit (Roche Sequencing Solutions, Pleasanton, CA, USA) following the manufacturer’s protocol, and lcWGS were subsequently performed following previously established protocols^1^. LcWGS data were used for copy number profiling with TFx estimation to identify recurrent genomic alterations in BC tissue samples. One tissue sample was discarded because it had fewer than 20M reads. CNV analyses were performed for 20 tissue samples using the ichorCNA algorithm^2^ with default settings, except for the following parameter modifications: normal = “c(0.2, 0.3, 0.4, 0.5, 0.6, 0.7, 0.8, 0.9)”, maxCN = 5, estimateScPrevalence = TRUE, scStates = “c(1,3)”, txnStrength = 10000. The panel of normals was generated from the plasma cfDNA samples from cancer-free controls. A TFx threshold of 0.03 was applied as the limit of detection for positive tumor signal detection.

#### Statistical analyses and data visualization

All statistical analyses were performed in R (v4.4.3)^3^, and visualizations were generated using R package ggplot2 (v.4.0.1)^4,5^. Continuous variables were compared using the Wilcoxon rank-sum test or Kruskal-Wallis test with Dunn’s post hoc test, as appropriate. P values were adjusted for multiple testing using the Benjamini-Hochberg method, with statistical significance defined as adjusted p < 0.05. Median cumulative cfDNA fragmentation distributions were compared using the Kolmogorov–Smirnov test.

## Supplementary Figures

**Supplementary Figure S1:**
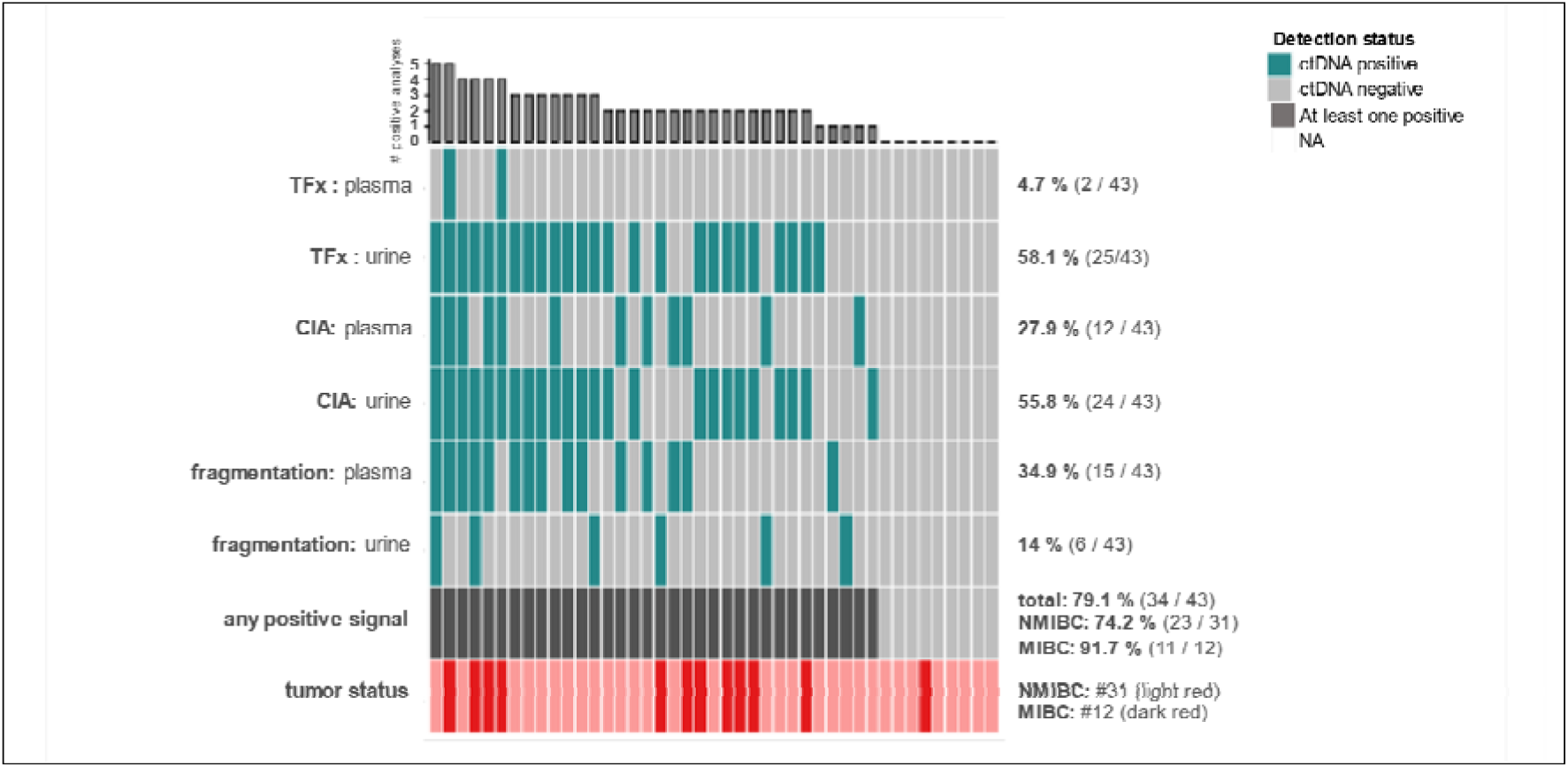
Multimodal ctDNA detection in plasma and urinary cfDNA from the subset of BC patients with matched plasma-urine pairs. Oncoprint summarizing genomic (estimated TFx based on CNVs, CIA score) and fragmentomic (plasma 10-bp oscillation score, urinary P163–169 bp) analyses of plasma and urinary cfDNA from 43 BC patients with matched plasma-urine pairs. Colored tiles indicate ctDNA-positive results in the respective analyses. The bar plot above the oncoprint shows the number of positive analyses per patient. The second-to-last row represents the number of BC patients with at least one ctDNA-positive result. Abbreviations: cfDNA, cell-free DNA; CIA, chromosomal instability analysis; CNV, copy number variation; ctDNA, circulating tumor DNA; LBx, liquid biopsy; MIBC, muscle-invasive bladder cancer; NMIBC, non-muscle-invasive bladder cancer; P163–169 bp, proportion of cfDNA fragments with lengths of 163–169 bp; TFx, tumor fraction.

**Supplementary Figure S2:**
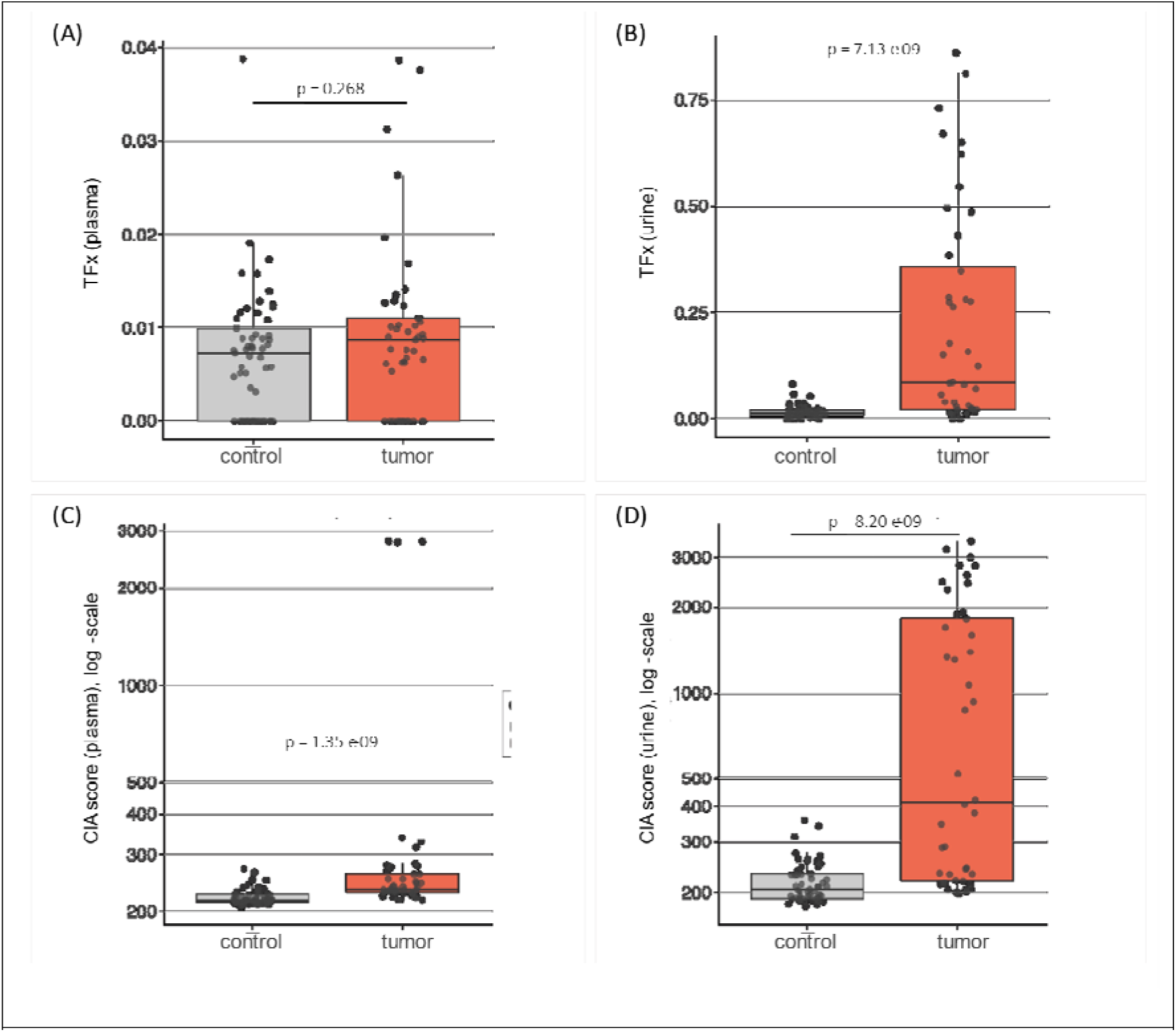
CNV and chromosomal instability profiling in plasma and urinary cfDNA. (A+B) Comparison of TFx values in plasma cfDNA (A) or urinary cfDNA (B) across cancer-free controls and BC patients. **(C+D)** Comparison of CIA scores in plasma cfDNA (C) or urinary cfDNA (D) across cancer-free controls and BC patients. **(A–D)** Box plot center lines indicate the median, and boxes illustrate the interquartile range with Tukey whiskers. Each dot represents one sample. Groups were compared using Wilcoxon testing.

**Supplementary Figure S3:**
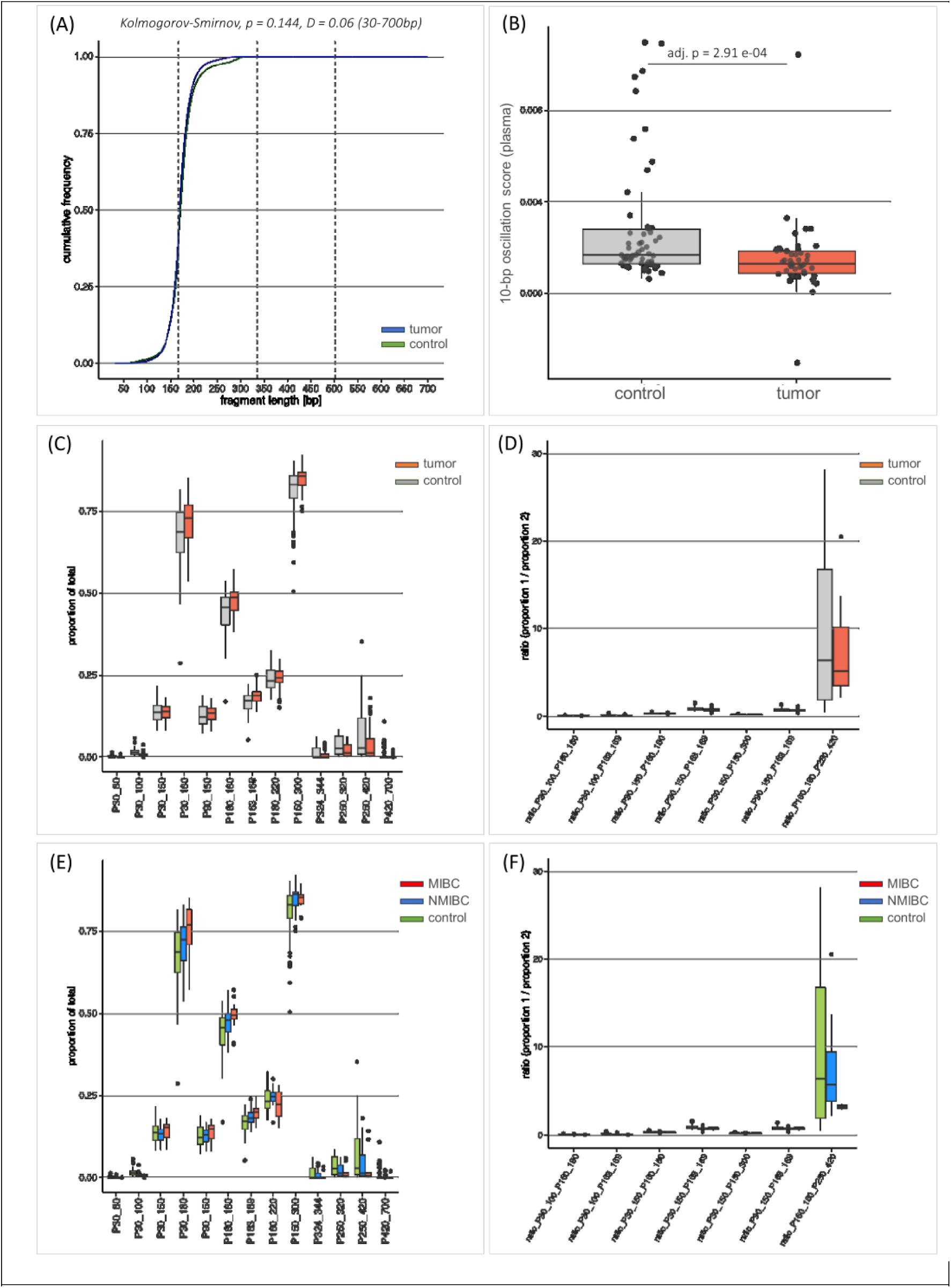
Plasma cfDNA fragmentation profiling. **(A)** Median cumulative fragment length distributions of plasma cfDNA from cancer-free controls, NMIBC patients, and MIBC patients. Y-axis: Cumulative frequency of cfDNA fragments of a given length (bp) normalized to all fragments between 30 and 700 bp. Vertical gray dotted line(s) indicate 167 bp and its multiples, 334 bp (2 x 167 bp) and 501 bp (3 x 167 bp). Median cumulative distributions of all BC patients and cancer-free controls were compared using the Kolmogorov-Smirnov test. **(B)** Comparison of plasma 10-bp oscillation scores between cancer-free controls and BC patients. Box plot center lines indicate the median, and boxes illustrate the interquartile range with Tukey whiskers. Each dot represents one sample. Groups were compared using Wilcoxon testing. **(C)** Comparison of proportions of plasma cfDNA fragment length ranges between BC patients and cancer-free controls. **(D)** Comparison of ratios of fragment length range proportions between BC patients and cancer-free controls. **(E)** Comparison of proportions of plasma cfDNA fragment length ranges across cancer-free controls, NMIBC patients, and MIBC patients. **(F)** Comparison of ratios of fragment length range proportions across cancer-free controls, NMIBC patients, and MIBC patients. **(C–F)** Box plot center lines indicate the median, and boxes illustrate the interquartile range with Tukey whiskers. Dots represent outliers. Abbreviations. D, Kolmogorov-Smirnov test D statistic

**Supplementary Figure S4:**
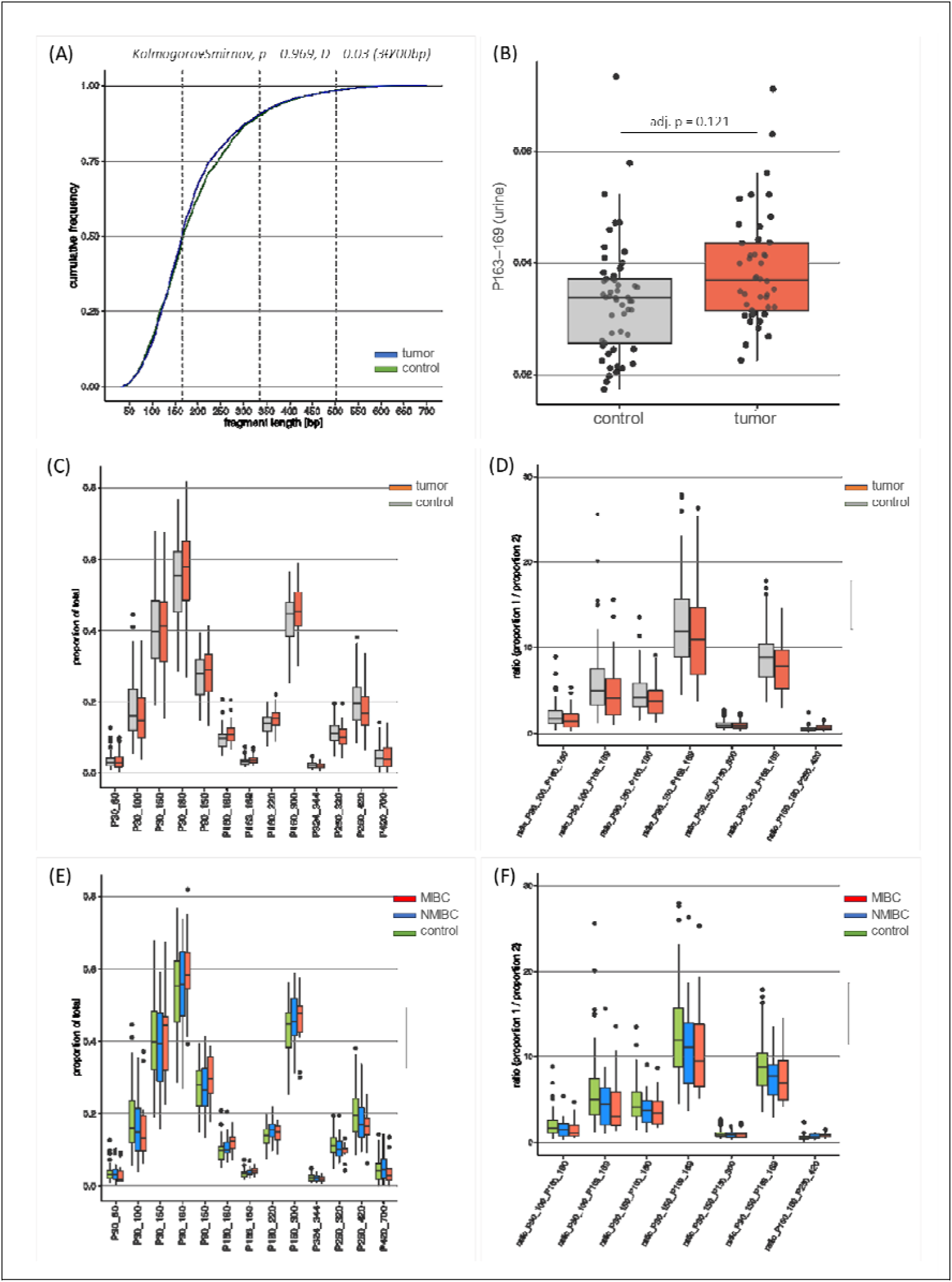
Urinary cfDNA fragmentation profiling. **(A)** Median cumulative fragment length distributions of urinary cfDNA from cancer-free controls, NMIBC patients, and MIBC patients. Y-axis: Cumulative frequency of cfDNA fragments of a given length (bp) normalized to all fragments between 30 x 167 bp) and 501 bp (3 x 167 bp). Median cumulative distributions of all BC patients and cancer-free controls were compared using the Kolmogorov-Smirnov test. **(B)** Comparison of urinary P163–169 bp values between cancer-free controls and BC patients. Box plot center lines indicate the median, and boxes illustrate the interquartile range with Tukey whiskers. Each dot represents one sample. Groups were compared using Wilcoxon testing. **(C)** Comparison of proportions of urinary cfDNA fragment length ranges between BC patients and cancer-free controls. **(D)** Comparison of ratios of fragment length range proportions between BC patients and cancer-free controls. **(E)** Comparison of proportions of urinary cfDNA fragment length ranges across cancer-free controls, NMIBC patients, and MIBC patients. **(F)** Comparison of ratios of fragment length range proportions across cancer-free controls, NMIBC patients, and MIBC patients. **(C–F)** Box plot center lines indicate the median, and boxes illustrate the interquartile range with Tukey whiskers. Dots represent outliers.

